# Community pharmacists’ referrals to General Practice with suspected need of antibiotics: a national prospective pilot

**DOI:** 10.1101/2024.06.19.24309200

**Authors:** Paulina Stehlik, Rebekah Moles, Mark Jones, Amanda Murray, Sarira El-Den, Mark Morgan, Chris Del Mar

**Author notes:** Corresponding author Dr Paulina Stehlik. Data availability statement All additional data and analysis code can be accessed via our OSF page (https://osf.io/2yngt/) or by contacting the lead author. Author contributions Paulina Stehlik and Chris Del Mar conceptualized the project. Paulina Stehlik, Chris Del Mar, Mark Morgan, Rebekah Moles, Mark Jones and Ian Fredericks contributed to the methodology. Paulina Stehlik, Chris Del Mar, Rebekah Moles, and Ian Fredericks acquired funding. Paulina Stehlik developed the research data base. Amanda Murry, Paulina Stehlik, Mark Morgan, Sarira El-Den and Rebekah Moles all contributed to recruitment and Amanda Murry acted as project coordinator and conducted data entry. Analysis was performed by Paulina Stehlik and Mark Jones. Paulina Stehlik and Chris Del Mar write the original draft, all authors reviewed and edited the final manuscript.

## Abstract

**Background:** Interventions to minimise community antibiotic use have focused on the GP and patient behaviour rather than the community pharmacist. Patient expectations are a known driver for antibiotic prescribing, and pharmacists may be inveterately contributing to these expectations by referring patients for GPs for suspected antibiotic-requiring infections (S-ARI). We sought to quantify these referral rates.

**Method:** Pharmacists and GPs were recruited independently using convenience sampling and completed prospective data collection on 20 minor ailment encounters and consecutive consultations respectively. Pharmacists recorded patient gender, age, referral reason and comments (if any). GPs recorded patient age, gender, reason for visit, and origin of patient referral including self-referral.

All data were analysed descriptively. Generalised estimating equations, multivariable logistic regression was used to investigate factors that may be associated with pharmacist referral rates.

**Results:** Nineteen pharmacists representing 466 minor ailments encounters, and 19 GPs representing 394 consultations were recruited.

Pharmacists referred 16.5% (77/466) of all minor ailments encounters for S-ARI. Comments suggested that reasons included upper-respiratory tract, ear nose and throat, and urinary tract infections. Most of S-ARI referrals were to a GP (62/466).

None of the 88 consultations for infection in GP data were documented as being referred by a pharmacist; majority were self-referred (77.3%; 68/88).

**Discussion:** Pharmacists referred 1 in 8 minor ailments encounters to the GP for S-ARI, with some indication they were for conditions that do not require antibiotics. Most GP consultations for infection were documented as self-referrals. Both provide potential points of intervention to minimise antibiotic use.

## Background

Antibiotic resistance is a global public health concern and is predicted to cause 10 million deaths annually by 2050.(1) In Australia, antibiotic resistance directly results in approximately 1600 deaths a year.(2) Despite current efforts to minimise use and curb antibiotic resistance, Australia still has one of the highest rates of community antibiotic prescribing in the world,(3) prescribing 4-9 times higher than recommended.(4) Most antimicrobial prescriptions in community settings are written by general practitioners (GPs), and The Australian Commission on Safety and Quality in Health care AURA 2021 found very high rates of inappropriate antibiotic prescribing for conditions with no evidence of benefit – for example over 80% of patients were prescribed an antibiotic for acute bronchitis.(5)

There are several reasons that GPs may overprescribe antibiotics including fear of financial loss, or overestimation of prescription effectiveness.(6) More importantly, time-poor GPs may believe it is quicker to complete the consultation by issuing a prescription, particularly if there is a perception that the patient expects an antibiotic and non-prescription may jeopardise the GP-patient relationship.(6)

While most interventions aiming to minimise unnecessary antibiotic use in community have targeted GPs and patients, the role of the community pharmacists (henceforth called pharmacists) has often been forgotten.(6) This is despite estimates suggesting that patients see their pharmacist 1.5-10 times more than their GP(7), with patients in Australia seeing pharmacists 18 times a year on average.(8) Patients often visit pharmacies to check whether they should see the GP,(9) and approximately 5% of all Australian GP visits are a result of pharmacist referral.(10) However, the rate of referral from pharmacist to GP varies from study to study and reasons for referrals have not been systematically categorised.(10-14) Nonetheless, studies that have looked at overall general requests in Australia and the US demonstrate that between 0.4-4% of all minor ailment visits to pharmacies resulted in referral,(13, 14) and can be as high as 94% when patients present with specific problems.(11)

What is not known, is how frequently pharmacists refer to the GP for suspected antibiotic requiring infections (S-ARI). Given that this may potentially contribute to unnecessary visits and patient expectations, quantifying the extent of referral for antibiotics is warranted.

The primary aim of this pilot study was to evaluate the feasibility of a national survey of pharmacist referrals for suspected need of antibiotics, specifically recruitment, survey usability and provide information for a more robust sample size calculation. The secondary aim was to quantify pharmacist referral rates to GPs for antibiotics and identify areas for further exploration.

## Methods

### Study design & Setting

We used a cross-sectional study design. Pharmacists and GPs were asked to prospectively fill out a paper-based post-consultation record form (supplementary files 1 and 2) for 20 consecutive patients. For pharmacists this meant 20 consecutive minor ailment patient encounters, and for GPs this meant 20 consecutive GP consultations.

Ethics approval was provided by Bond University Human Research Ethics Committee (PS00123).

### Participants

All registered pharmacists working in community pharmacy and medical practitioners working in general practice in Australia were eligible to participate in the study. Other staff, such as pharmacy students, interns, technicians, and assistants, were not eligible to participate in the study.

### Recruitment

We used purposive and convenience sampling methods for recruitment from May to December 2019. Pharmacists and GPs were recruited independently of one another through practice-relevant magazines and newsletters, as well as through social media, colleagues and known contacts. This was supplemented by snowballing. Emails and social media posts were sent out three times per recruitment round at weeks 0, 2 and 6. We repeated this process one month after week 6 throughout the recruitment period.

### Data Collection

We used two separate but analogous paper-based post-consultation record forms for pharmacists and GPs.

Pharmacists were asked to complete the pharmacist post-consultation record form documenting information about the patient, and if, how, why and to whom they referred the patient. Additional information collected included: pharmacy type and location, busyness, and date and time of data collection. The full data collection form and definitions are provided in supplementary file 1.

GPs were asked to complete the GP post-consultation record form documenting information about the patient and presenting problem. GPs were also required to ask their patients whether they had been referred and to record the source of referral for each consultation. Additional information collected included clinic type, GP location, and month of data collection. The full data collection form and definitions are provided in supplementary file 2.

### Changes to original protocol

#### Recruitment

We made two separate amendments for additional recruitment strategies. The first was made after we had recruited 15 GPs and 4 pharmacists (approved 14 Aug 2019) to allow for recruitment via students attending their clinical placements who were asked to pass study information on to their preceptors and were allowed to facilitate data entry under preceptor instruction. This resulted in one additional GP and 5 additional pharmacists recruited. The second amendment was to allow for direct approach of GPs and pharmacists via phone or face-to-face (approved 6 Nov 2019) by the research team and resulted in the remaining GP and pharmacist recruitments.

#### Data collection

We assumed that multiple pharmacists would contribute to data collection and initially asked pharmacists to complete and collect data on 50 minor ailment consultations. Feedback indicated that this took too long as data was collected on their own. We changed the form from 50 to 20 patients after receiving two forms back.

### Sample size

#### Community pharmacists

We piloted our data collection tool with two Victorian, one NSW and one Queensland pharmacists. Data indicated pharmacists dealt with approximately 10 minor ailments per day, and 10% of these were referred to a GP for antibiotics. Assuming that it would take approximately one week to have 50 minor ailment encounters, we required a minimum of 10 pharmacies to estimate the percentage of consultations that result in referral to a GP for antibiotics to within +/- 5% with 95% confidence.

#### General practitioners

We piloted our data collection tool with two Queensland GPs. They used the tool after 36 patient consultations and found none were referred by a pharmacist and documented 33% of consultations for infection. Assuming GPs provide approximately 30 consultations a day (given a 15 min time(15)), we required a minimum of 21 GPs to collect data about at least 20 patient consultations to estimate the percentage of patients referred to a GP by a pharmacist that were to within +/- 5% with 95% confidence.

### Statistical methods

Descriptive statistics were used to analyse data. Missing data were reported as a category. Analysis was conducted in R 3.6.3 using DescTools package. Data were wrangled into an analysable form in Python 3.8.5 using pandas, numpy, datetime, and xlrd packages.

We conducted an exploratory analysis in SAS 9.4 for Windows to describe the potential impact of variables on pharmacist referral rates. The unit of analysis was consultations for minor ailments within pharmacies. Generalized estimating equations multivariable logistic regression were used to investigate factors that may be associated with referral for any reason, and referral for antibiotic. The factors were gender, age group, rurality and how busy the pharmacist was. An exchangeable correlation structure was specified to allow for within pharmacy correlations. This analysis was purely exploratory and hypothesis-generating in nature intended to inform future study design.

Due to a lack of pharmacist referrals for antibiotics identified in GP data, we did not conduct an exploratory analysis of the GP data.

## Results

### Feasibility

We had difficulties in recruitment which resulted in several changes to our original protocol (see above). Overall, we found that directly approaching/cold calling the most successful recruitment strategy for pharmacists, and personal contacts and professional social media advertisements for GPs.

Early on we noticed that we were having difficulty in getting data from pharmacists (see above) resulting in a change in the data collection from 50 to 20 minor ailment which improved recruitment. Feedback from pharmacist and GP participants was that the form was quick and easy to use.

Feedback from participants who had expressed interest but were then unwilling to participate indicated that the 2019-2020 bushfires and then subsequently the start of the COVID19 pandemic were significant barriers to study participation. We stopped data collection as participants were unable to commit to participating due to the COVID19 pandemic.

### Participants

A total 50 pharmacists and 48 GPs agreed to participate; 19 GP and 19 pharmacist forms were returned. Data were collected from May 2019 until Jan 2020.

### Community Pharmacy Data

Of the 19 pharmacies, 13 (68%) were chain pharmacies, 16/19 (84%) were in urban areas and 9/19 (47%) were in NSW and 4/19 (21%) in Queensland. Fifteen (79%) forms were completed by pharmacists working as the sole pharmacist, and 11/19 (58%) pharmacies described “average” busyness during data collection. Data were collected across office and out-of-office hours, predominantly during the spring and summer months, and it took an average of two days to complete data collection.

Pharmacists collected data on 466 patients presenting with minor ailments; 79% came in for themselves, 55% were female, and 72% were aged 13-65 (Tables

Table 1).

**Table 1:** Community Pharmacy Results.

| Name | n | % (95% CI) |
| --- | --- | --- |
| Number | 466<sup>1</sup> | - |
| Referred | 173/466 | 37 (33, 42) |
| **Why was the patient referred?** |  |  |
| S-ARI<sup>2</sup> | 77/466 | 17 (12, 21) |
| Reason other than S-ARI | 88/466 | 19 (15, 23) |
| Not reported | 8/466 | 2 (0, 6) |
| **For all minor ailments referred for S-ARI: What was the type of referral?** |  |  |
| Conditional<sup>3</sup> | 12/466 | 3 (0, 6) |
| Direct<sup>4</sup> | 61/466 | 13 (10, 17) |
| Not reported | 4/466 | 1 (0, 4) |
| **Antibiotic referral: Who was the patient referred to?** |  |  |
| GP | 62/466 | 13 (10, 17) |
| ER | 2/466 | 0.4 (0, 4) |
| Other HP | 2/466 | 0.4 (0, 4) |
| Not reported | 11/466 | 2 (0, 6) |

<sup>1</sup> Includes 2 pharmacies that collected on 20 minor ailment encounters; remaining 17 pharmacies collected on 20 pharmacies only.
<sup>2</sup> S-ARI: Suspected antibiotic requiring infection. Pharmacists documented a referral as S-ARI if they suspected that the patient had an infection requiring antibiotic treatment, even if they did not explicitly tell the patient.
<sup>3</sup> Direct referral: Patient was asked to see medical practitioner ASAP.
<sup>4</sup> Conditional referral: Patient was asked to see medical practitioner if certain criteria are met. For example, if symptoms don't resolve within a given time frame, or if additional symptoms appear.

Pharmacists referred 173/466 (37%) minor ailment patients for medical review, and 77/466 (17%) were for S-ARI. Of patients referred for S-ARI, 62/466 (13%) patients were referred to a GP, with 46/466 (9%) recommended to go directly to the GP, 12/466 (3%) conditionally (if certain criteria/symptoms met), and 1 did not have the ‘type’ of referral recorded (Tables Table 1).

Some pharmacists left comments for the reason for referral for 40 instances where patients were referred for S-ARI. Most were for suspected skin and soft tissue infections (10 instances) and unspecified upper respiratory tract infections (7 instances) (Table 2).

**Table 2.**
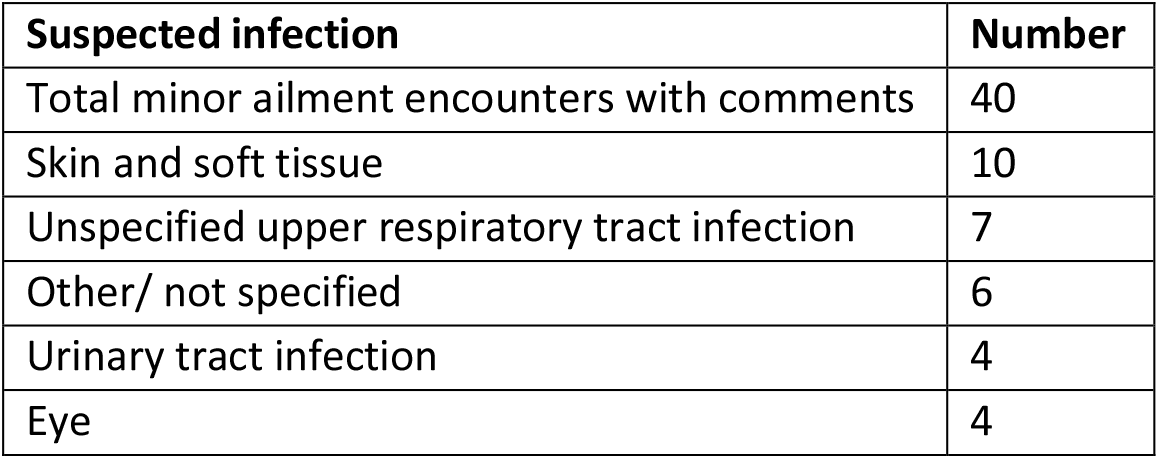
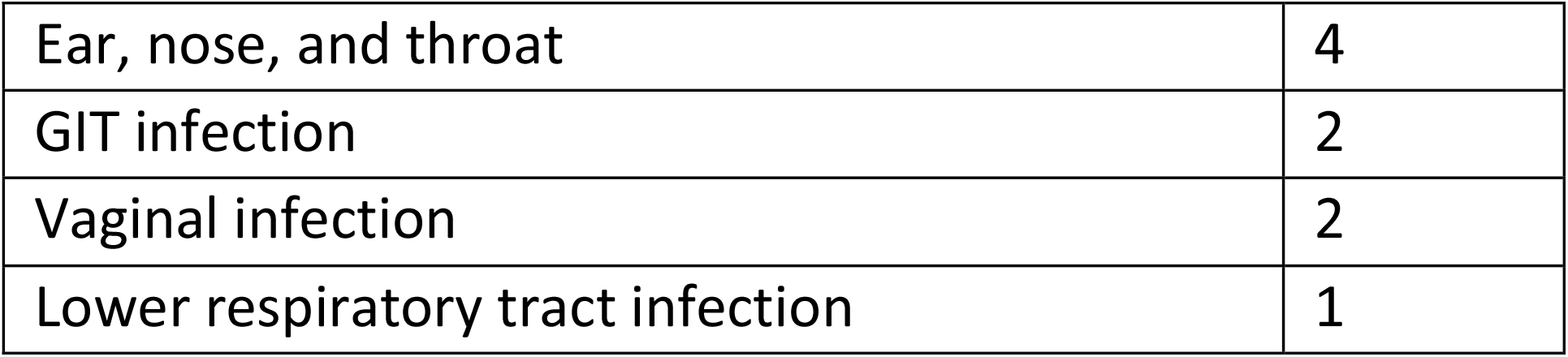
S-ARI reason for referrals provided by pharmacist in free text comments.

**Table 3:** General practice results.

| Name | n | % (95% CI) |
| --- | --- | --- |
| Number | 394 | - |
| **Reason for visit** |  |  |
| Infection | 88/394 | 22 (18, 26) |
| Medication related problem | 45/394 | 11 (9, 15) |
| Other | 283/394 | 72 (68, 76) |
| Not reported | 6/394 | 2 (0.5, 3) |
| **Infection related visit: Who was documented as the referring person?** |  |  |
| Pharmacist | 0/88 | 0 (0, 2) |
| Self | 68/88 | 77 (69, 86) |
| Family | 17/88 | 19.3 (13, 28) |
| Other | 3/88 | 3 (1, 7) |
| Not reported | 1/88 | 1 (0, 3) |

Pharmacists were less likely to refer patients in older age groups for antibiotics and were more likely to refer patients for any reason if they were from rural areas and when the pharmacy was busier (Supplementary Tables 1 and 2).

### General Practice Data

Of the 19 GP practices, 13/19 (68%) were traditional GP practices, 10/19 (53%) bulk-billed, 11/19 (58%) co-located with a pharmacy, 12/19 (64%) were in urban areas, and 15/19 (79%) were in Queensland. Sixteen (84%) GPs collected data during office hours, and mostly during the winter months.

GPs collected data on 394 consultations; 57% of patients were female, and 57% aged 13-65 (Table).

Of the 88/394 (22%) consultations for infection, none were documented as being referred by a pharmacist. Most were documented as self-referrals (68/88; 77%) (Table 5).

Overall, there were only four documented referrals by a pharmacist (1%). All were for medication-related problem consultations.

## Discussion

Our study indicates that that measuring pharmacist referral rates for S-ARI is feasible with a simple data collection tool. It also provides information for a more robust sample size calculation and study design for a national level study and has identified several areas for further investigation.

We found that pharmacists referred 1 in 8 of all minor ailment encounters were to a GP for S-ARI. This was not reflected in GP data where only 4 consultations were recorded as having been referred by a pharmacist, none of which were for an infection-related consultation. Almost all GP consultations for infection were documented as either self-referred or referred by a family member. This number is significantly lower than previous findings in Australia.(10) While reasons for these differences were not explored, one explanation might be that less than 50% of patients follow pharmacist referral advice due to the informal nature of the encounter.(11, 16)

Our results suggest opportunities for new interventions to minimise antibiotic use by minimising unnecessary GP visits for antibiotics. We estimate that, given that approximately 22% of all GP consultations were for infection, and 20-51% of all patients referred by pharmacists eventually go to the GP,(11) pharmacists could be contributing 4-12% of all GP visits to GPs for antibiotics. While the appropriateness of referral was not examined in this study, comments provided by pharmacists suggest that they referred patients for S-ARI for conditions for which there is little to no benefit of antibiotic benefit – namely unspecified upper respiratory tract infections, eye infections, ear nose and throat infections, and urinary tract infections.(5)

The high number of self- and family-referral rates for infection consultations in the GP data suggest that pharmacist triage could also be leveraged to minimise GP visits for self-limiting conditions such as acute respiratory infections, and to manage patient expectations for antibiotics through the use of shared-decision making. Formal minor ailment schemes in the UK and Canada have been successful at doing this(17) and in reducing primary health care costs.(18) No such scheme exists in Australia but, like other countries, could be used as part of a national anti-microbial stewardship strategy.(19)

This study has several strengths. Firstly, GPs and pharmacists collected data prospectively minimising the potential for recall bias. Secondly, pharmacists completed the data collection form rather than using observational techniques. Previous studies have used observation, exit interviews, or mystery shopper techniques to evaluate pharmacy triage(11); however, this does not capture internal intent for referral that may not be made explicit to the patient.

Nonetheless, results should be interpreted with caution as this was a pilot study with a small sample size and may have overestimated referral rates. We found higher referral rates compared to earlier studies (up to 15%),(11, 16) although some studies have estimated pharmacists refer up to 94% of patients depending on the presenting condition.(11) Our sample also over-represented urban areas and eastern states and may not reflect other areas of Australia. Some of this may be explained by difficulties experienced with recruitment and return of data collection forms. Early on this was due to the number of patients we had asked pharmacists to collect data about; changing from 50 to 20 minor ailment encounters improved recruitment rates. Later difficulty in recruitment and form return may have been related to the 2019 Australian summer bushfires and then the start of the COVID19 pandemic. As recruitment strategies were implemented in stages, it appeared that the email of close contacts and social media and cold calls were most effective, and recruitment via students was least effective.

## Conclusions

This pilot study found that pharmacists often refer patients to GPs for antibiotics, but that GPs may be unaware of this. The reason for this discrepancy warrants further exploration. Our data also suggests that pharmacists may be referring patients for S-ARI for conditions where there is little to no evidence that antibiotics are needed. The findings of this study can be used to develop a national survey to further explore the appropriateness of pharmacist referrals and identify potential interventions for pharmacists to minimise unnecessary antibiotic use.

## Data Availability

All data produced in the present study are available upon reasonable request to the authors.

## Acknowledgements

We would like to acknowledge Ian Fredericks for his input during the design phase of this project. We would also like to thank Dr Mina Bakhit for his constructive feedback on our manuscript draft.

## Supplementary Tables

*Supp Table 1: Odds ratios for association between factors of interest and referral for S-ARI by Community Pharmacists*

*Supp Table 2: Odds ratios for association between factors of interest and referral for any reason by Community Pharmacists*

Supplementary File 1: Community Pharmacist Data Collection Form

Supplementary File 2: General Practice Data Collection Form

