## supplementary Table 1 for "Community pharmacists’ referrals to General Practice with suspected need of antibiotics: a national prospective pilot"

Supplementary Tables

*Supp Table 1: Odds ratios for association between factors of interest and referral for S-ARI by Community Pharmacists*

| **Factor** | **Odds ratio** | **95% CI** | **P-value** |
| --- | --- | --- | --- |
| Female | 0.77 | 0.43 – 1.39 | 0.40 |
| Increasing Age | 0.67 | 0.49 – 0.90 | 0.008 |
| Increasing Rurality | 1.29 | 0.98 – 1.71 | 0.066 |
| Increasingly Busy workplace | 1.71 | 0.96 – 3.03 | 0.066 |
