## supplementary Table 2 for "Community pharmacists’ referrals to General Practice with suspected need of antibiotics: a national prospective pilot"

Supplementary Tables

Supp Table 2: Odds ratios for association between factors of interest and referral for any reason by Community Pharmacists

| **Factor** | **Odds ratio** | **95% CI** | **P-value** |
| --- | --- | --- | --- |
| Female | 0.72 | 0.50 – 1.03 | 0.075 |
| Increasing Age | 1.04 | 0.72 – 1.52 | 0.82 |
| Increasing Rurality | 1.64 | 1.09 – 2.47 | 0.018 |
| Increasingly Busy workplace | 1.60 | 1.08 – 2.38 | 0.019 |
