## supplementary file 1 for "Community pharmacists’ referrals to General Practice with suspected need of antibiotics: a national prospective pilot"

### Supplementary File 1: Community Pharmacist record form

Dear Pharmacists

Please document the number of referrals **you make** for the **next** **20** **consecutive** **minor-ailments consultations (over approx. 1 working day).**

Only **fully registered pharmacists** that are involved in the minor ailment consultation may contribute to the data collection form. Student and intern pharmacists, and other pharmacy staff are NOT able to contribute.

##### Definitions:

**Minor-ailments consultation**: a consultation with a patient or carer regarding any symptom and product-based OTC requests that may or may not have resulted in the provision of a product and/or referral.

For example: management of upper respiratory tract infection, urinary tract infection, sty, provision of S3s, wound care, sun care, etc.

Note that a minor ailment is NOT a direct result of drug treatment (i.e. things NOT related issues to a prescription or adverse drug effect, or services provided) and do NOT include provision of pharmacy services such as NDSS and immunisation.

**Who was the patient?**

**Self**: if the person requesting the minor-ailment consultation for *themselves*.

**Proxy**: if the person requesting the minor-ailment consultation for *someone else*.

**If both**: please complete a separate entry for each person.

**Was a medical referral recommended?**

**No - referral not required**: Patient was not asked to see a medical practitioner directly or conditionally; other management strategies may have been offered (OTC, life style advice, non-pharmacological management, other counselling, etc). If a patient does not require a referral, you will only need to tick the minor ailment counter box, but you will not need to document a reason or where the patient was referred to.

**Yes - Direct referral**: Patient asked to see medical practitioner ASAP

**Yes - Conditional referral**: Patient asked to see medical practitioner *if* certain criteria are met. For example, if symptoms don’t resolve within a given time frame, or if additional symptoms appear.

**Reason for referral:** While pharmacists are not diagnosticians, community pharmacists frequently refer patients to medical practitioners when they suspect they have a diagnosis requiring a prescription, or when they are unsure of the diagnosis and feel the patient may need further medical examination.

**Patient likely needs ANTIBIOTICS**: if you suspect that the patient has an infection requiring antibiotic treatment, even if you did not explicitly tell the patient.

**Patient likely needs further medical evaluation and/or other prescription but NOT antibiotics**: if you suspect that the patient has a condition that may require further medical evaluation and that most likely not require antibiotics; for example, a GI bleed, or elevated blood pressure.

**Start date of data collection period: ____/___/2019	End date of data collection period: ____/___/2019**

Please complete the form below after for the next **20 consecutive minor ailment consultations** (tick all that apply):

| Patient number (office use) | Who was the patient? | |  | Patient Gender (M/ F/ O = Unknown/ other) | Approximate age of patient in YEARS:  < 2, 2-12, 13-65, > 65 |  | Was a medical referral recommended? | | |  | Reason for referral | |  | Where was the patient referred? | | |  | Comments |
| --- | --- | --- | --- | --- | --- | --- | --- | --- | --- | --- | --- | --- | --- | --- | --- | --- | --- | --- |
|  | Self | Proxy |  |  |  |  |  |  |  |  |  |  |  |  |  |  |  |  |
|  |  |  |  |  |  |  | **NO** - referral not required.  **NO MORE DATA TO RECORD.** | **Yes -** Direct | **Yes** - Conditional  If **Yes**, continue |  | Patient likely needs **ANTIBIOTICS** | Other - Something else but **NOT antibiotics** |  | General Practitioner | Emergency Department | Other |  |  |
| 1 |  |  |  |  |  |  |  |  |  |  |  |  |  |  |  |  |  |  |
| 2 |  |  |  |  |  |  |  |  |  |  |  |  |  |  |  |  |  |  |
| 3 |  |  |  |  |  |  |  |  |  |  |  |  |  |  |  |  |  |  |
| 4 |  |  |  |  |  |  |  |  |  |  |  |  |  |  |  |  |  |  |
| 5 |  |  |  |  |  |  |  |  |  |  |  |  |  |  |  |  |  |  |
| 6 |  |  |  |  |  |  |  |  |  |  |  |  |  |  |  |  |  |  |
| 7 |  |  |  |  |  |  |  |  |  |  |  |  |  |  |  |  |  |  |
| 8 |  |  |  |  |  |  |  |  |  |  |  |  |  |  |  |  |  |  |
| 9 |  |  |  |  |  |  |  |  |  |  |  |  |  |  |  |  |  |  |
| 10 |  |  |  |  |  |  |  |  |  |  |  |  |  |  |  |  |  |  |
| 11 |  |  |  |  |  |  |  |  |  |  |  |  |  |  |  |  |  |  |
| 12 |  |  |  |  |  |  |  |  |  |  |  |  |  |  |  |  |  |  |
| 13 |  |  |  |  |  |  |  |  |  |  |  |  |  |  |  |  |  |  |
| 14 |  |  |  |  |  |  |  |  |  |  |  |  |  |  |  |  |  |  |
| 15 |  |  |  |  |  |  |  |  |  |  |  |  |  |  |  |  |  |  |
| 16 |  |  |  |  |  |  |  |  |  |  |  |  |  |  |  |  |  |  |
| 17 |  |  |  |  |  |  |  |  |  |  |  |  |  |  |  |  |  |  |
| 18 |  |  |  |  |  |  |  |  |  |  |  |  |  |  |  |  |  |  |
| 19 |  |  |  |  |  |  |  |  |  |  |  |  |  |  |  |  |  |  |
| 20 |  |  |  |  |  |  |  |  |  |  |  |  |  |  |  |  |  |  |
| 21 |  |  |  |  |  |  |  |  |  |  |  |  |  |  |  |  |  |  |
| 22 |  |  |  |  |  |  |  |  |  |  |  |  |  |  |  |  |  |  |
| 23 |  |  |  |  |  |  |  |  |  |  |  |  |  |  |  |  |  |  |
| 24 |  |  |  |  |  |  |  |  |  |  |  |  |  |  |  |  |  |  |
| 25 |  |  |  |  |  |  |  |  |  |  |  |  |  |  |  |  |  |  |

**CONTINUED OVERLEAF…**

### Additional information

| About the Data collection period  With respect to the pharmacists collecting data, the pharmacists **mainly** worked:   - as the sole pharmacist - with other pharmacist(s)   With respect to the data collection period, was data collected during (tick all the apply):   - Weekday: normal office hours (Mon-Fri, 9-5) - Weekday: *outside* of normal office hours - Weekend   On average, how “busy” was the pharmacy during the data collection period?   - Very busy - Busier than usual - Average - Quiet - Very quiet | About the pharmacy |
| --- | --- |
|  | Pharmacy Type - Chain pharmacy - Independent pharmacy Pharmacy location information: Pharmacy postcode: _____ State or territory: - Qld \| - Vic \| - NSW \| - Tas; - ACT \| - NT \| - WA \| Region: - Urban \| - Regional; - Rural \| - Remote Which best describes the location of the pharmacy: - Next to/ collocated with GP Clinic - Medical centre - Hospital - Street: shopping-strip - Street: non-shopping strip - In a shopping centre - Other: ______________ |

**END OF SURVEY**

After you have completed the data collection sheet, please email to the study coordinator:
