## supplementary file 2 for "Community pharmacists’ referrals to General Practice with suspected need of antibiotics: a national prospective pilot"

### Supplementary file 2: General Practitioner record form

Dear General Practitioner

Please use the form below after **next** **20 consecutive** **consultations.**

##### Definitions:

**The Problem**: What did the patient suspect or what were they concerned about?

**Infection**: For any of the problems raised during the consultation, did the patient indicate that they suspected, or were concerned about, an *infection of any cause* (bacterial, viral, fungal, parasite or otherwise)?

**Medication related problem**: For any of the problems raised during the consultation, did the patient indicate that they suspected, or were concerned about, a *medication related problem* (adverse event, dose, interaction, drug choice, or otherwise)?

**Other**: For any of the problems raised during the consultation, where they related to *other* reasons (non-infection, non-medication related problems such as chronic disease state management, immunisation, check-up, administrative, etc)?

**Referral:** You will need to ask the patient if anyone had recommended that they visit you for any of the problems raised during the consultation:

Suggested wording: “*Did someone advise you or suggest that you come in about any of your concerns?*”

**Self**: the patient was not referred by anyone other than themselves.

**Family member or friend** recommended that they attend for any one of the problems raised during the consultation.

**Pharmacist** recommended that they attend for any one of the problems raised during the consultation.

**Other:** any other person who may recommended that they attend for any one of the problems raised during the consultation.

Please complete the form below after for the next **20 consecutive consultations** (tick all that apply):

| Patient number (office use) | Patient | |  | **The Problem**: What did the **patient** suspect or were concerned about? | | |  | Referred by: | | | |  | Comments |
| --- | --- | --- | --- | --- | --- | --- | --- | --- | --- | --- | --- | --- | --- |
|  | Age:  < 2, 1-12, 12-65, > 65 | Patient Gender (M/ F/ O = Unknown/ other) |  | Infection? | Medication related problem? | Other: chronic disease management, checkup, administrative, etc. |  | Self | Family member or friend | Pharmacist | Other |  |  |
| 1 |  |  |  |  |  |  |  |  |  |  |  |  |  |
| 2 |  |  |  |  |  |  |  |  |  |  |  |  |  |
| 3 |  |  |  |  |  |  |  |  |  |  |  |  |  |
| 4 |  |  |  |  |  |  |  |  |  |  |  |  |  |
| 5 |  |  |  |  |  |  |  |  |  |  |  |  |  |
| 6 |  |  |  |  |  |  |  |  |  |  |  |  |  |
| 7 |  |  |  |  |  |  |  |  |  |  |  |  |  |
| 8 |  |  |  |  |  |  |  |  |  |  |  |  |  |
| 9 |  |  |  |  |  |  |  |  |  |  |  |  |  |
| 10 |  |  |  |  |  |  |  |  |  |  |  |  |  |
| 11 |  |  |  |  |  |  |  |  |  |  |  |  |  |
| 12 |  |  |  |  |  |  |  |  |  |  |  |  |  |
| 13 |  |  |  |  |  |  |  |  |  |  |  |  |  |
| 14 |  |  |  |  |  |  |  |  |  |  |  |  |  |
| 15 |  |  |  |  |  |  |  |  |  |  |  |  |  |
| 16 |  |  |  |  |  |  |  |  |  |  |  |  |  |
| 17 |  |  |  |  |  |  |  |  |  |  |  |  |  |
| 18 |  |  |  |  |  |  |  |  |  |  |  |  |  |
| 19 |  |  |  |  |  |  |  |  |  |  |  |  |  |
| 20 |  |  |  |  |  |  |  |  |  |  |  |  |  |
| 21 |  |  |  |  |  |  |  |  |  |  |  |  |  |
| 22 |  |  |  |  |  |  |  |  |  |  |  |  |  |
| 23 |  |  |  |  |  |  |  |  |  |  |  |  |  |
| 24 |  |  |  |  |  |  |  |  |  |  |  |  |  |
| 25 |  |  |  |  |  |  |  |  |  |  |  |  |  |

Data collection dates occurred during which month(s) (complete as many as required):

| **Month:** | - May | - June | - July | - Aug | - Sept | - Oct | - Nov |
| --- | --- | --- | --- | --- | --- | --- | --- |

PTO

### Additional information

About the Data collection period

With respect to the data collection period, was data collected during (tick all the apply):

- Weekday: normal office hours (Mon-Fri, 9-5)
- Weekday: *outside* of normal office hours
- Weekend

About the GP Clinic

Clinic Type

- Traditional GP Practice
- Medical Centre
- Specialist Clinic

Is the GP clinic:

- Bulk-bill
- Private

cLINIC location information:

Clinic postcode: _____

State or territory:

| - Qld | - Vic | - NSW | - Tas |
| --- | --- | --- | --- |
| - ACT | - NT | - WA |  |

Region:

| - Urban | - Regional |
| --- | --- |
| - Rural | - Remote |

Is the GP clinic **co-located with** or **next to a pharmacy**?

- Yes
- No

**END OF SURVEY**

After you have completed the data collection sheet, please email to the study coordinator:
